# Border-Region Status and Diagnosed Diabetes Prevalence in Texas: A Cross-Sectional Ecological Analysis

**DOI:** 10.64898/2026.05.30.26354501

**Authors:** Priyanka Rani Saha, Sagor Khan, Yussif Yahaya, Md Al Amin Meia

## Abstract

Diagnosed diabetes disproportionately burdens socioeconomically disadvantaged populations in the United States, particularly Hispanic communities in the Texas–Mexico border region. Few studies have quantified whether geographic border-region status is independently associated with county-level diagnosed diabetes prevalence after accounting for lifestyle and food-environment factors. This cross-sectional ecological study examined 253 Texas counties using CDC PLACES 2025 health estimates and USDA Food Environment Atlas food-access data, including the 2015 county-level low-food-access measure. Border-region counties were defined using the official La Paz Agreement 32-county definition, which includes counties within 100 km of the US–Mexico boundary. Multiple linear regression with HC3 robust standard errors was used to estimate associations between border-region status, low food access, physical inactivity, and diagnosed diabetes prevalence. Variance inflation factor analysis assessed multicollinearity, and Global Moran’s *I* tested spatial autocorrelation in diagnosed diabetes prevalence and OLS residuals. Border-region counties had 33% higher unadjusted mean diagnosed diabetes prevalence than non-border counties (16.1% vs. 12.1%). After adjustment, border-region status remained significantly associated with a 0.625 percentage-point higher diagnosed diabetes prevalence (*β* = 0.625, 95% CI [0.357, 0.893], *p <* 0.001). Physical inactivity was the strongest independent predictor (*β* = 0.404, 95% CI [0.391, 0.417], *p <* 0.001). The model explained 96.0% of county-level variance (*R*^2^ = 0.960, *N* = 253), reflecting ecological associations among modeled county-level health indicators. Global Moran’s *I* confirmed strong spatial clustering of diagnosed diabetes prevalence (*I* = 0.5734, *p* = 0.001), with reduced but significant residual spatial autocorrelation after OLS adjustment (*I* = 0.1696, *p* = 0.001). These findings suggest that border-region status is associated with elevated diagnosed diabetes prevalence beyond physical inactivity and low food access, supporting targeted public health investment in the Texas–Mexico border region.

## 1 Introduction

Diagnosed diabetes mellitus affects approximately 38 million Americans and represents a substantial public health challenge in the United States [1]. The burden of this disease is not uniformly distributed across the population; rather, it disproportionately affects Hispanic and Latino populations, low-income communities, and geographically isolated regions [2, 3]. Hispanic adults, in particular, bear a significant burden, with type 2 diabetes prevalence approximately 70% higher than that of non-Hispanic white adults [4].

The Texas–Mexico border region offers a critical case study for investigating these health disparities. The La Paz Agreement of 1983 defined the border area as the 100 km (62.1 mile) buffer zone on each side of the international boundary, a definition that remains a standard geographic unit for border health research and policy [5, 6]. This region, particularly the Rio Grande Valley, comprising Hidalgo, Cameron, Starr, and Willacy counties, is characterized by a predominantly Hispanic population facing systemic socioeconomic challenges, including high poverty rates and limited healthcare access [7, 8]. Localized studies in Cameron County have documented diabetes prevalence exceeding 30% in some communities, emphasizing the need for broader statewide analysis [9].

Food access is an important structural determinant of metabolic health. The United States Department of Agriculture (USDA) defines low food access in relation to household income and distance from supermarkets or grocery stores [10]. Limited access to nutritious food has been linked to poor dietary quality, obesity, and metabolic disease burden [11, 12]. Previous county-level ecological studies have demonstrated associations between food environment variables and diabetes prevalence [13, 14]; however, the combined role of food access and border-region geography in Texas has not been fully characterized.

Physical inactivity is also one of the strongest modifiable risk factors for type 2 diabetes [15]. Population-level inactivity is strongly correlated with county-level diabetes burden [16], but existing research in Texas has often relied on descriptive comparisons, localized border-community studies, or simple correlation methods [17]. There remains limited evidence regarding whether border-region county status is independently associated with diagnosed diabetes prevalence after accounting for county-level physical inactivity and food access.

This study addresses this gap by applying multiple linear regression with heteroskedasticity-consistent standard errors to county-level data from 253 Texas counties with available information. Using the formal La Paz Agreement definition as a binary geographic co-variate, we tested the hypothesis that Texas–Mexico border-region county status is significantly associated with diagnosed diabetes prevalence after controlling for county-level physical inactivity and low food access.

## 2 Methods

### 2.1 Study Design and Setting

This cross-sectional ecological study utilized the county as the primary unit of analysis. The study population comprised 253 of the 254 counties in Texas. Loving County was excluded from the analysis due to the suppression of diabetes prevalence data in federal datasets, resulting in a final analytical sample of *N* = 253. As the study relied exclusively on publicly available, de-identified, county-level aggregate data, it did not constitute human subjects research, and institutional review board approval was not required.

### 2.2 Data Sources

#### 2.2.1 CDC PLACES 2025

County-level health estimates were retrieved from the Centers for Disease Control and Prevention (CDC) PLACES: Local Data for Better Health, 2025 release [18]. PLACES utilizes a multilevel regression and poststratification (MRP) framework to link Behavioral Risk Factor Surveillance System (BRFSS) survey responses with high-resolution demographic and socioeconomic data from the US Census Bureau’s American Community Survey. This methodology generates validated model-based age-adjusted prevalence estimates for all US counties [19, 20]. Because these estimates are produced through a standardized small-area modeling framework rather than direct county-level sampling alone, associations among PLACES-derived health indicators should be interpreted as ecological relationships between modeled population estimates rather than as individual-level associations.

#### 2.2.2 USDA Food Environment Atlas 2025

Environmental and food access indicators were obtained from the US Department of Agriculture (USDA) Economic Research Service Food Environment Atlas, 2025 release [10]. Although the Atlas was released in 2025, specific food-access metrics, including low access to grocery stores, reflect 2015 data, which remains the most recent available county-level release for these specific indicators. These measures were used as proxies for relatively stable structural characteristics of the county-level built environment. Because diagnosed diabetes prevalence reflects chronic disease burden shaped by long-term social, behavioral, and environmental exposures, the 2015 food-access measure provides relevant historical context for interpreting current county-level diabetes patterns. Nevertheless, the temporal gap between the food-access measure and the 2025 health estimates is acknowledged as a limitation.

### 2.3 Variables

#### 2.3.1 Outcome Variable

The primary outcome was the county-level age-adjusted prevalence of diagnosed diabetes among adults (%). This measure represents the percentage of the adult population that has been told by a health professional that they have diabetes, excluding gestational diabetes. The measure does not distinguish between Type 1 and Type 2 diabetes.

#### 2.3.2 Primary Predictor (Border-Region Status)

Border-region status was operationalized as a binary geographic covariate (1 = border, 0 = non-border). Following the legal definition established by the 1983 La Paz Agreement, border counties were defined as those situated within 100 km (62.1 miles) of the US–Mexico international boundary [5]. This encompasses 32 Texas counties, as recognized by the US–Mexico Border Health Commission [6]. A complete list of all 32 border-region counties is provided in Supplementary Table S4. Sensitivity analysis using a narrower 20-county border definition is reported in Supplementary Table S2.

#### 2.3.3 Covariates

Two primary structural and behavioral covariates were included in the final regression model:

i. **Low food access:** The percentage of the county population with low access to grocery stores, defined using USDA Food Environment Atlas criteria based on distance from supermarkets or large grocery stores in urban and rural areas [10].
ii. **Physical inactivity:** The age-adjusted prevalence (%) of adults aged *≥* 18 years who reported no leisure-time physical activity in the past 30 days [18].

Obesity prevalence was assessed descriptively and in multicollinearity diagnostics but was excluded from the final regression model because of extreme multicollinearity with physical inactivity (VIF *>* 100).

### 2.4 Statistical Framework

Descriptive statistics were computed for all variables, stratified by border-region status. Because both the outcome variable and key lifestyle covariates were derived from the CDC PLACES multilevel regression and poststratification framework, the regression results were interpreted as ecological associations among modeled county-level health indicators. This methodological alignment may contribute to a high coefficient of determination (*R*^2^), because the model captures structured relationships among pre-modeled population estimates rather than the greater stochastic variation expected in individual-level survey data. The primary association was estimated using a multiple linear regression model:

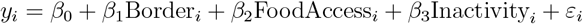

where *y*_*i*_ represents the age-adjusted diagnosed diabetes prevalence for county *i*. Prior to estimation, the Breusch–Pagan test was conducted, which indicated significant heteroskedasticity (*χ*^2^ = 11.20, *p* = 0.047). Consequently, the model was estimated using an HC3 heteroskedasticity-consistent covariance matrix. HC3 was selected over the more common HC0 estimator because simulation evidence suggests that HC3 provides more reliable heteroskedasticity-consistent inference in finite samples, particularly when leverage points are present [21].

### 2.5 Spatial Autocorrelation Framework

Given the geographic nature of county-level data, we formally tested for spatial dependence in both diagnosed diabetes prevalence and OLS model residuals using Global Moran’s *I*. A Queen’s contiguity spatial weights matrix was constructed using the `libpysal` package [22]. Counties were defined as neighbors if they shared any common boundary point or vertex. The matrix was row-standardized so that each row summed to unity. The final spatial weights matrix included all 253 counties, with no isolated counties. Global Moran’s *I* was calculated for both age-adjusted diagnosed diabetes prevalence and OLS model residuals as:

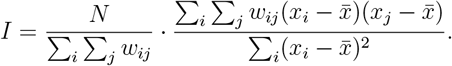

where *N* is the number of counties (*N* = 253), *w*_*ij*_ is the row-standardized spatial weight between counties *i* and *j* from the Queen’s contiguity matrix, *x*_*i*_ is the value of interest for county *i*, either age-adjusted diagnosed diabetes prevalence or OLS residuals, and 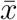 is the mean across all counties. The expected value under spatial randomness is:

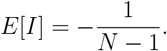

Statistical significance was assessed using 999 conditional permutations. Sensitivity analysis examined whether findings were robust to alternative border county definitions. Statistical significance for regression and correlation analyses was defined as *p <* 0.05.

### 2.6 Computational Environment

All primary regression analyses were conducted in Python 3.9 using a local Jupyter Notebook environment. Spatial autocorrelation diagnostics were performed using Python on the UTRGV CRADLE high-performance computing cluster. The final analysis used `statsmodels` 0.14.6, `scipy` 1.14.1, `libpysal` 4.13.0, and `esda` 2.7.0. Analysis code and the final dataset are publicly available, as described in the Data Availability statement.

## 3 Results

### 3.1 Descriptive Statistics

Of 254 Texas counties, 253 had complete data and were included in the analysis. Thirty-two counties were classified as border-region counties and 221 as non-border counties under the La Paz Agreement definition. Table 1 presents descriptive statistics stratified by border-region county status.

**Table 1.** Descriptive statistics by border-region county status, Texas counties (N=253).

| Variable | Border-region<br>(n=32) | Non-border<br>(n=221) | All counties<br>(n=253) |
| --- | --- | --- | --- |
| Diagnosed diabetes (%) | 16.1 (10.4–21.5) | 12.1 (8.7–17.5) | 12.6 (8.7–21.5) |
| Obesity (%) | 41.0 (33.6–48.2) | 37.3 (29.4–43.9) | 37.7 (29.4–48.2) |
| Physical inactivity (%) | 37.6 (23.8–48.9) | 29.2 (19.9–41.6) | 30.2 (19.9–48.9) |
| Low food access (% , 2015) | 43.9 (7.8–100.0) | 29.3 (0.0–100.0) | 31.1 (0.0–100.0) |

*Note.* Values are means with ranges shown in parentheses. Border-region counties were defined using the 32-county La Paz Agreement definition, corresponding to counties within 100 km of the US–Mexico boundary. Low food access reflects 2015 USDA Food Environment Atlas data. Dimmit County had the highest diagnosed diabetes prevalence in the analytical dataset (21.5%). Real and Terrell counties recorded complete low food access exposure (100.0%) among La Paz border-region counties, while Hudspeth County also showed very high low food access exposure (99.6%). The US national average diagnosed diabetes prevalence was 12.7%. Data sources: CDC PLACES 2025; USDA Food Environment Atlas 2025.

Border-region counties had substantially higher mean diagnosed diabetes prevalence (16.1% vs. 12.1%), obesity prevalence (41.0% vs. 37.3%), physical inactivity (37.6% vs. 29.2%), and low food access exposure (43.9% vs. 29.3%) compared to non-border counties. As illustrated in Fig. 1, border-region counties showed higher medians and wider distributions for the descriptive indicators shown in the box plots, with statistically significant differences confirmed by Mann–Whitney U tests (*p <* 0.001).

**Fig 1.**
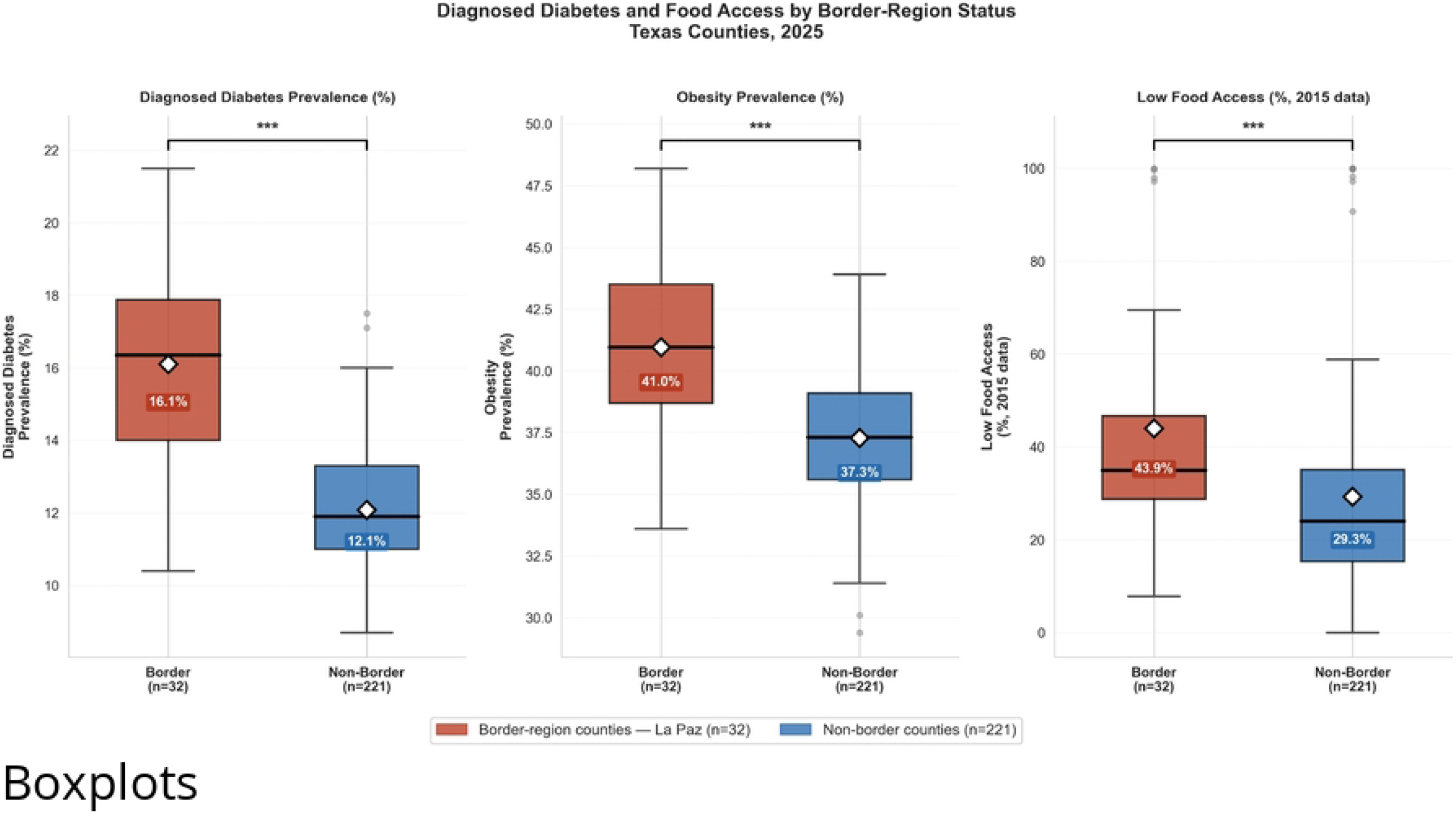
Descriptive comparison of diagnosed diabetes, obesity, and low food access between La Paz border-region and non-border Texas counties. Box plots comparing (A) diagnosed diabetes prevalence, (B) obesity prevalence, and (C) low food access exposure using 2015 USDA Food Environment Atlas data between La Paz border-region counties (dark red, n=32) and non-border counties (blue, n=221). Diamond markers indicate group means. Border-region counties show higher median values and wider distributions across all three indicators. Significance brackets reflect Mann– Whitney U tests. *** *p <* 0.001. Data sources: CDC PLACES 2025; USDA Food Environment Atlas 2025.

Dimmit County recorded the highest diagnosed diabetes prevalence in the analytical dataset (21.5%), followed by Zavala County (21.4%) and Starr County (20.5%), all well above the US national average of 12.7%. Real and Terrell counties recorded complete low food access exposure (100.0%) among La Paz border-region counties, while Hudspeth County also showed very high low food access exposure (99.6%). As shown in Fig. 2, 20 of the top 25 highest-diabetes counties in Texas were border-region counties, with all 25 counties exceeding the US national average. The geographic concentration of this burden is further illustrated in Fig. 3, which shows spatial clustering of high-prevalence counties along the southern border region, largely within the La Paz boundary.

**Fig 2.**
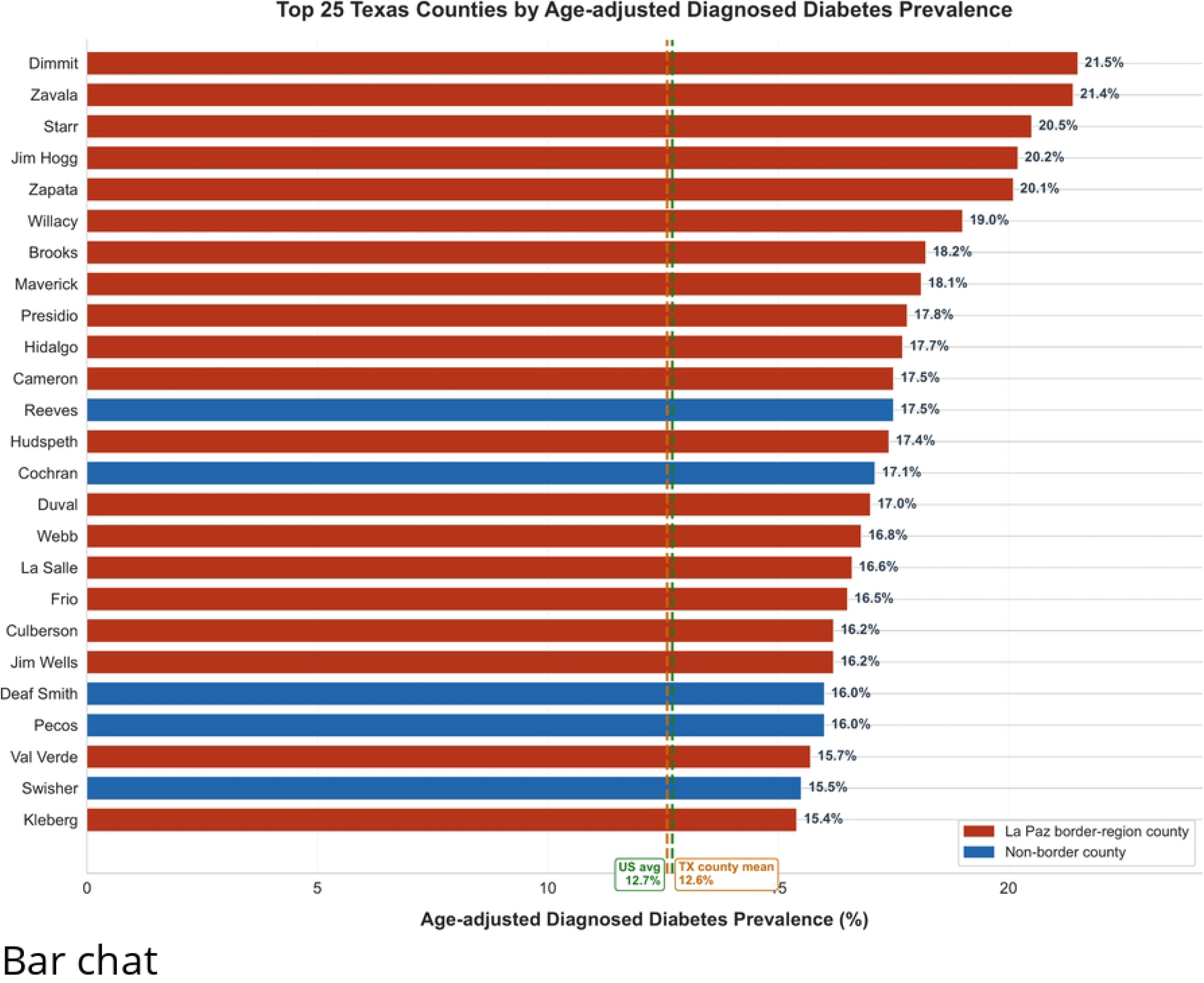
Top 25 Texas counties by age-adjusted diagnosed diabetes prevalence. Horizontal bar chart showing the 25 Texas counties with the highest age-adjusted diagnosed diabetes prevalence (%). Dark red bars indicate La Paz border-region counties, based on the 32-county La Paz Agreement definition, and blue bars indicate non-border counties. The vertical green dashed line indicates the US national average diagnosed diabetes prevalence (12.7%), while the vertical orange dashed line indicates the Texas county mean calculated from the analytical county dataset. Twenty of the top 25 counties with the highest diagnosed diabetes prevalence were classified as La Paz border-region counties. Data source: CDC PLACES 2025.

**Fig 3.**
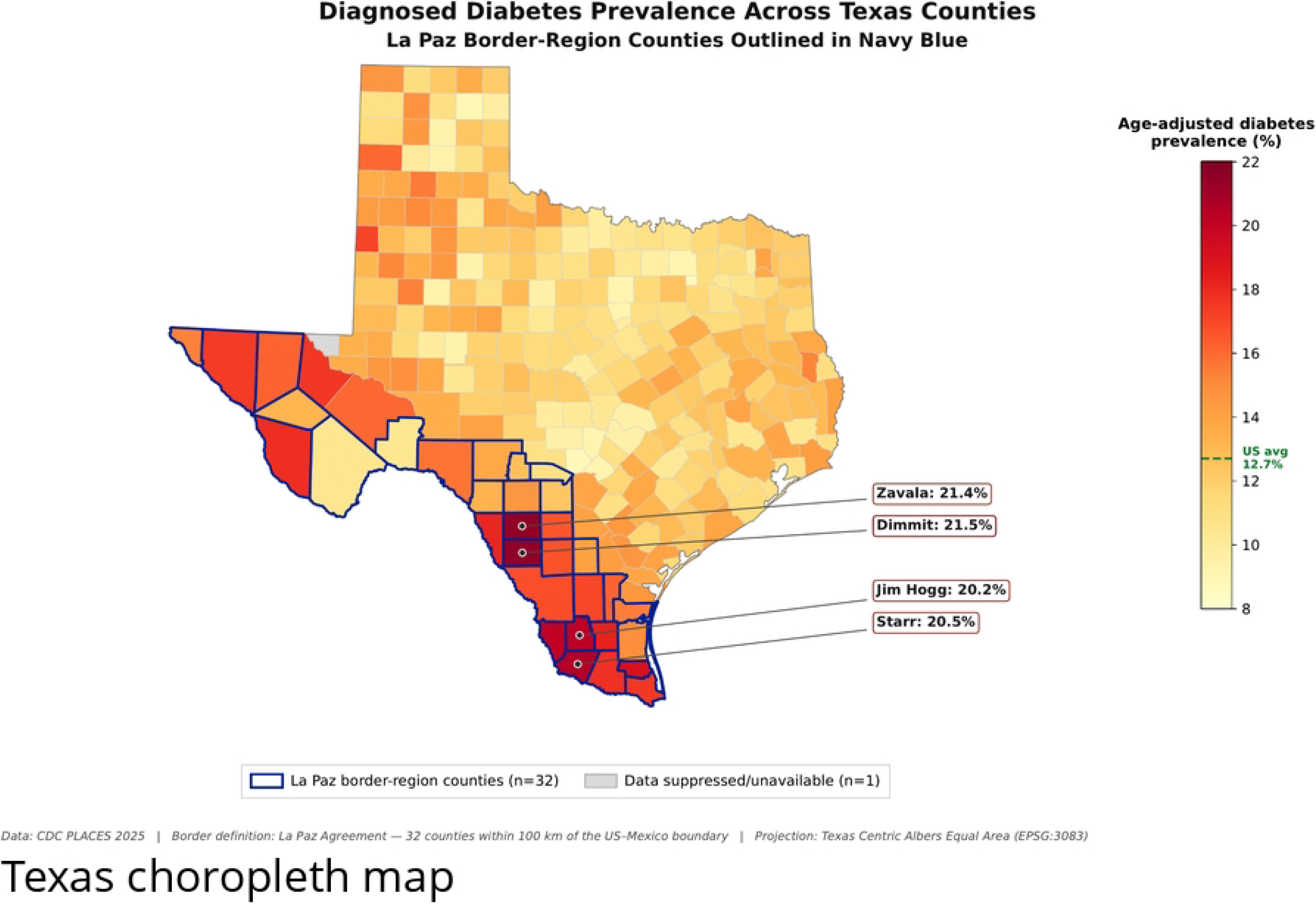
Geographic distribution of age-adjusted diagnosed diabetes prevalence across Texas counties, 2025. Choropleth map showing age-adjusted diagnosed diabetes prevalence (%) for the 253 Texas counties included in the analysis. The color scale ranges from light yellow, indicating lower prevalence, to dark red, indicating higher prevalence. Navy blue outlines indicate La Paz border-region counties (n=32), defined as counties within 100 km of the US–Mexico international boundary under the La Paz Agreement. The map shows a spatial concentration of high-burden counties along the southern Texas–Mexico border region, including Dimmit (21.5%), Zavala (21.4%), Starr (20.5%), and Jim Hogg (20.2%) counties. The green dashed line on the colorbar indicates the US national average diagnosed diabetes prevalence (12.7%). Gray shading indicates suppressed or unavailable county-level diabetes data (n=1). Projection: Texas Centric Albers Equal Area (EPSG:3083). Data sources: CDC PLACES 2025; county boundary file: US Census Bureau 2023 county cartographic boundary shapefile.

### 3.2 Correlation Analysis

Spearman correlation analysis revealed that physical inactivity (*ρ* = 0.972, *p <* 0.001) and obesity (*ρ* = 0.869, *p <* 0.001) were most strongly correlated with county-level diagnosed diabetes prevalence (Table 2). Border-region county status showed a significant positive correlation with diagnosed diabetes prevalence (*ρ* = 0.423, *p <* 0.001).

**Table 2.** Spearman correlation coefficients between diagnosed diabetes prevalence and predictor variables, Texas counties (N=253).

| Variable | $\rho$ | $p$-value | Significance |
| --- | --- | --- | --- |
| Physical inactivity (%) | 0.972 | $p < 0.001$ | *** |
| Obesity (%) | 0.869 | $p < 0.001$ | *** |
| Border-region county (La Paz, 1 = yes) | 0.423 | $p < 0.001$ | *** |
| No vehicle and low access (% , 2015) | 0.279 | $p < 0.001$ | *** |
| Low income and low access (% , 2015) | 0.236 | $p < 0.001$ | *** |
| Low food access (% , 2015) | 0.069 | $p = 0.275$ | ns |

*Note.* $\rho$ denotes the Spearman rank correlation coefficient. Statistical significance is indicated as \*\*\* $p < 0.001$ ; ns = not significant. Low food access variables reflect 2015 USDA Food Environment Atlas data. Data sources: CDC PLACES 2025; USDA Food Environment Atlas 2025.

The correlation heatmap (Fig. 4) reveals two distinct clusters of highly inter-correlated variables. The first cluster, comprising diagnosed diabetes, obesity, and physical inactivity, shows very strong mutual correlations (*ρ >* 0.87). The second cluster, comprising low food access, low income combined with low food access, and no vehicle access combined with low food access, shows strong mutual correlations (*ρ* = 0.91). Border-region county status shows moderate positive correlations with both clusters, supporting its interpretation as a distinct geographic and structural indicator rather than a proxy for lifestyle factors alone.

**Fig 4.**
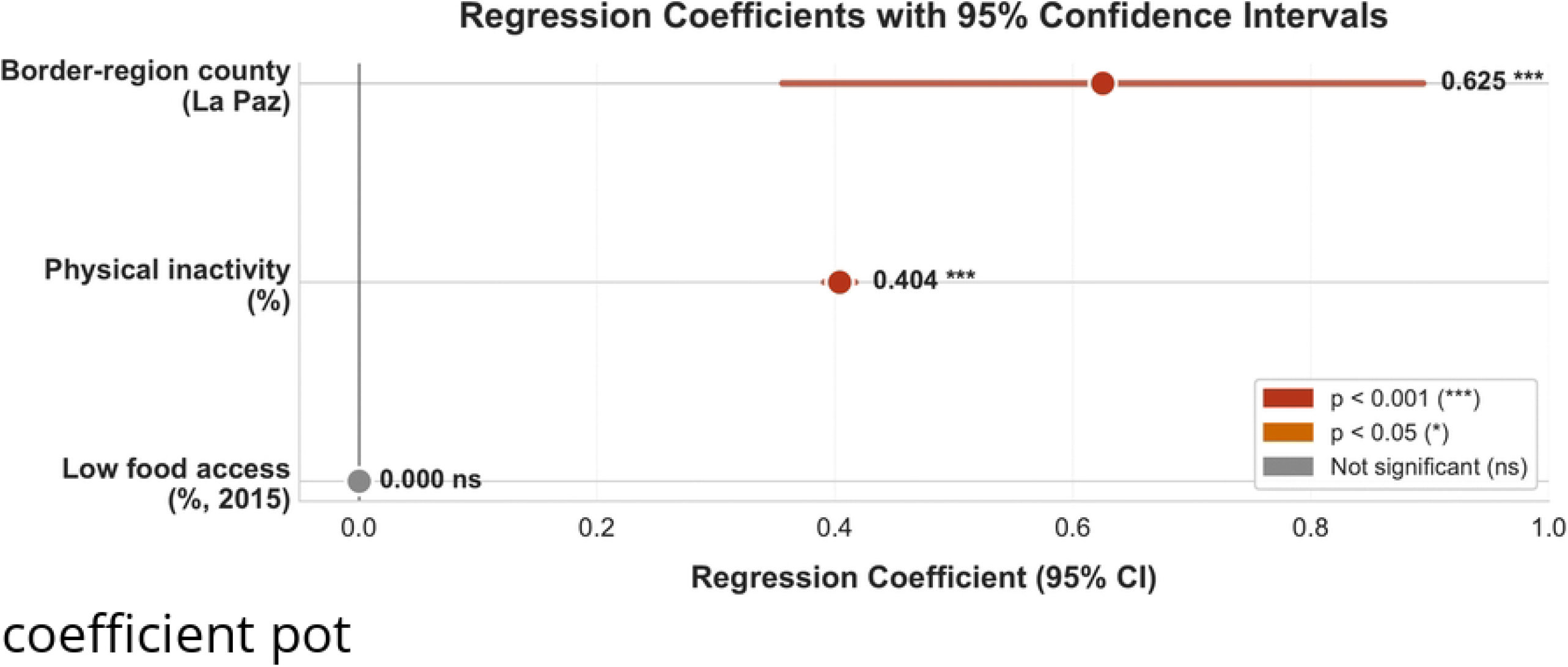
Spearman correlation matrix of diagnosed diabetes prevalence and associated factors, Texas counties (N=253). Heatmap showing Spearman rank correlation coefficients (*ρ*) among county-level diagnosed diabetes prevalence and selected predictor variables. Two major correlation patterns are visible: (1) a lifestyle-related cluster comprising diagnosed diabetes prevalence, obesity, and physical inactivity, with strong positive correlations among these variables (*ρ >* 0.87); and (2) a food-access cluster comprising low food access, low-income population with low food access, and no-vehicle households with low food access, with the strongest within-cluster correlation reaching *ρ* = 0.91. The color scale indicates positive correlations in red, near-zero correlations in white, and negative correlations in blue. Data sources: CDC PLACES 2025; USDA Food Environment Atlas 2025.

Notably, general low food access alone was not significantly associated with diagnosed diabetes prevalence (*ρ* = 0.069, *p* = 0.275), whereas the combination of low income and low food access (*ρ* = 0.236, *p <* 0.001) and absence of vehicle access combined with low food access (*ρ* = 0.279, *p <* 0.001) were both significant.

### 3.3 Multicollinearity Assessment

VIF analysis identified severe multicollinearity among obesity (VIF = 126.79), physical inactivity (VIF = 127.85), and the food access composite variables (VIF *>* 14). The reduced model retained physical inactivity (VIF = 2.71), low food access (VIF = 2.61), and border-region county status (VIF = 1.25), all well below the threshold of concern.

### 3.4 Regression Analysis

The VIF-corrected OLS regression with HC3 robust standard errors is presented in Table 3 and Fig. 5. The model explained 96.0% of county-level variance in diagnosed diabetes prevalence (*R*^2^ = 0.960, *F* = 1555.17, *p <* 0.001, *N* = 253). This high coefficient of determination should be interpreted as a methodological characteristic of the ecological and model-based data structure rather than as evidence of individual-level predictive certainty. Both the outcome variable and the physical inactivity covariate were derived from the CDC PLACES multilevel regression and poststratification framework; therefore, the regression model captures structured ecological relationships among modeled county-level health indicators. In this context, the high *R*^2^ reflects strong alignment among population-level modeled estimates rather than the independent stochastic variation typically observed in raw individual-level survey data.

**Table 3.** OLS regression with HC3 robust standard errors predicting county-level diagnosed diabetes prevalence, Texas counties (N=253).

| Variable | $\beta$ | Robust SE | 95% CI | $p$-value | Sig. |
| --- | --- | --- | --- | --- | --- |
| Intercept | 0.287 | 0.210 | [−0.124, 0.699] | 0.171 |  |
| Low food access (% , 2015) | 0.000 | 0.001 | [−0.002, 0.002] | 0.844 |  |
| Physical inactivity (%) | 0.404 | 0.007 | [0.391, 0.417] | < 0.001 | *** |
| Border-region county (La Paz) | 0.625 | 0.137 | [0.357, 0.893] | < 0.001 | *** |
| $R^2$ | 0.960 | | | | |
| Adjusted $R^2$ | 0.960 | | | | |
| F-statistic | 1555.17 ($p < 0.001$) | | | | |
| N counties | 253 |  |  |  |  |

*Note.* $\beta$ denotes the unstandardized regression coefficient. SE denotes HC3 heteroskedasticity-consistent robust standard error; CI denotes confidence interval. The outcome variable was age-adjusted diagnosed diabetes prevalence (%). Obesity and food access composite variables were excluded from the final model because of high variance inflation factors ( $VIF > 10$ ; see Supplementary Table S1). Breusch–Pagan heteroskedasticity test: $LM = 11.20$ , $p = 0.047$ . The high model fit ( $R^2 = 0.960$ ) should be interpreted cautiously because the outcome and physical inactivity predictor are both derived from the CDC PLACES multilevel regression and poststratification framework. \*\*\* $p < 0.001$ .

**Fig 5.**
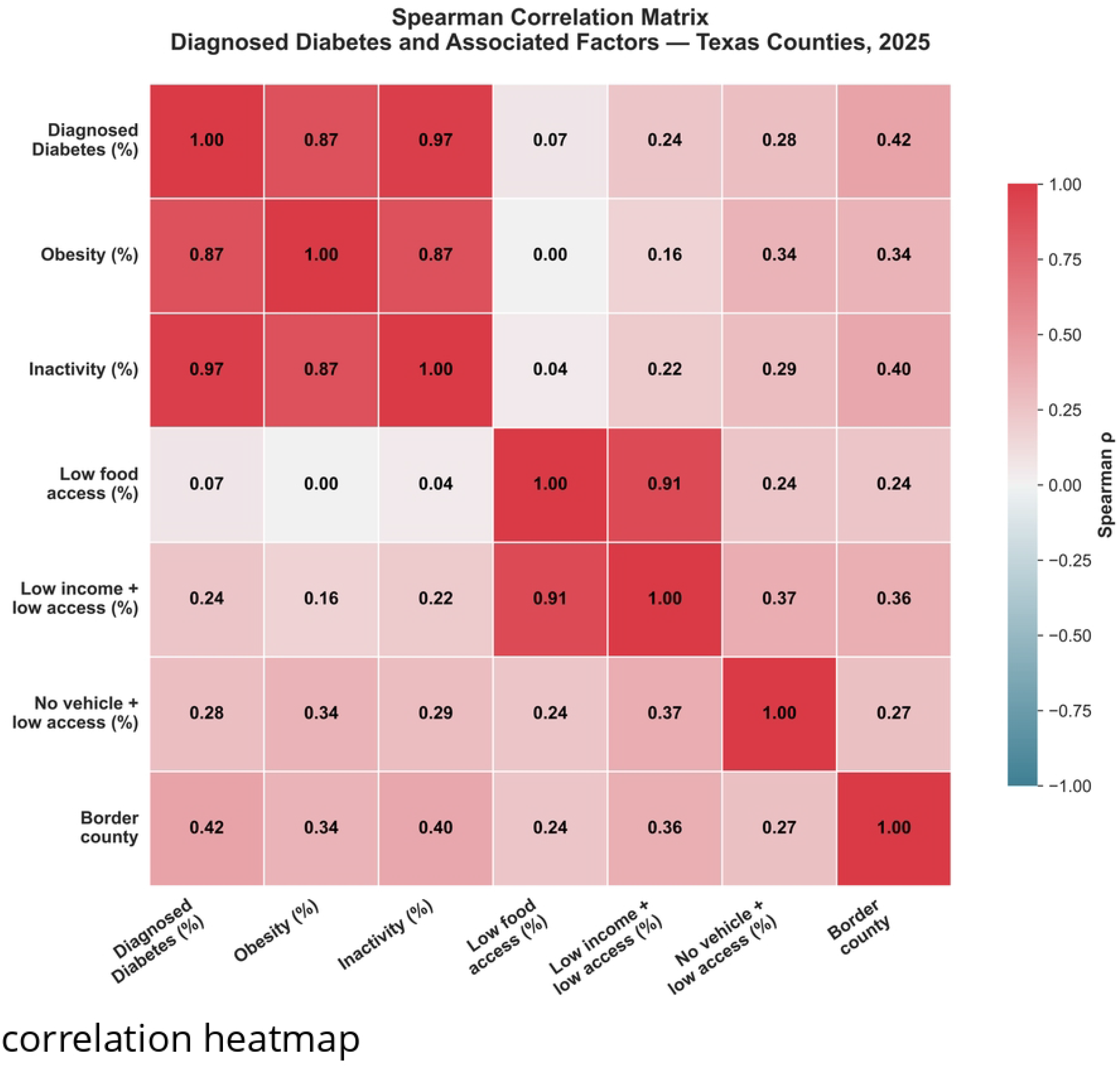
OLS regression coefficients with 95% confidence intervals for predictors of county-level diagnosed diabetes prevalence, Texas counties (N=253). Forest plot showing unstandardized OLS regression coefficients (*β*) with HC3 heteroskedasticity-consistent robust 95% confidence intervals [21]. Red markers indicate statistically significant predictors (*p <* 0.001), and gray markers indicate non-significant predictors. Border-region county status (*β* = 0.625) and physical inactivity (*β* = 0.404) both show confidence intervals entirely above zero, indicating statistically significant positive associations with diagnosed diabetes prevalence after adjustment. Low food access was not statistically significant in the adjusted model. Data sources: CDC PLACES 2025; USDA Food Environment Atlas 2025.

Physical inactivity was the strongest independent predictor (*β* = 0.404, SE = 0.007, 95% CI [0.391, 0.417], *p <* 0.001), indicating that each one percentage-point increase in physical inactivity was associated with a 0.404 percentage-point increase in diagnosed diabetes prevalence, controlling for other variables in the model.

Border-region county status remained a significant independent predictor after controlling for physical inactivity and low food access (*β* = 0.625, SE = 0.137, 95% CI [0.357, 0.893], *p <* 0.001). This adjusted association is shown in the coefficient plot (Fig. 5), while the scatter plot in Fig. 6 shows that several border-region counties were positioned above the fitted physical-inactivity trend line. Low food access alone was not a significant independent predictor (*β* = 0.000, *p* = 0.844).

**Fig 6.**
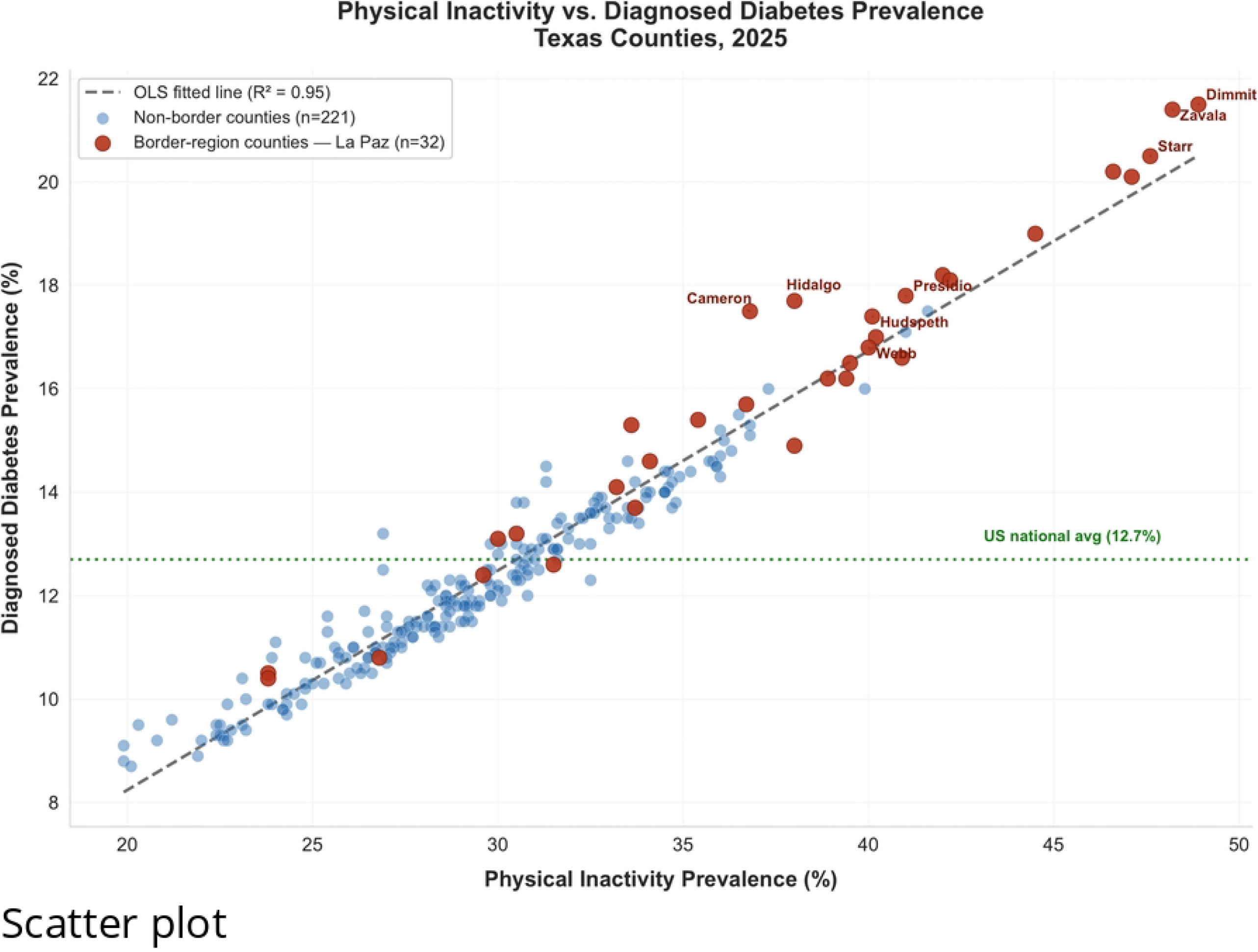
Physical inactivity versus diagnosed diabetes prevalence across Texas counties, stratified by border-region status. Scatter plot of county-level physical inactivity prevalence (%) versus age-adjusted diagnosed diabetes prevalence (%) for 253 Texas counties. Dark red circles indicate La Paz border-region counties (n=32), and blue circles indicate non-border counties (n=221). The dashed gray line represents the OLS fitted line (*R*^2^ = 0.95), and the green dotted line indicates the US national average diagnosed diabetes prevalence (12.7%). Several border-region counties appear above the fitted line, suggesting higher diagnosed diabetes prevalence than expected based on physical inactivity alone. Data sources: CDC PLACES 2025; USDA Food Environment Atlas 2025.

The coefficient plot in Fig. 5 confirms that border-region county status and physical inactivity were the only statistically significant predictors, with both confidence intervals entirely above zero.

### 3.5 Sensitivity Analysis

The border-region association was significant under both the narrower 20-county border definition (*β* = 1.004, 95% CI [0.703, 1.305], *p <* 0.001) and the 32-county La Paz definition (*β* = 0.625, 95% CI [0.357, 0.893], *p <* 0.001), confirming that findings were robust to border definition choice (Supplementary Table S2).

### 3.6 Spatial Autocorrelation Analysis

The Queen’s contiguity weights matrix included all 253 counties, with no islands and an average of 5.70 neighbors per county (range: 1–9). Global Moran’s *I* confirmed strong positive spatial clustering of diagnosed diabetes prevalence across Texas counties (*I* = 0.5734, *z* = 15.45, *p* = 0.001; Supplementary Fig. S2(A)), indicating that high-prevalence counties tended to cluster geographically near other high-prevalence counties. This finding is consistent with the geographic concentration observed in the choropleth map (Fig. 3).

Moran’s *I* applied to OLS model residuals indicated significant but substantially reduced residual spatial autocorrelation (*I* = 0.1696, *z* = 4.64, *p* = 0.001; Supplementary Fig. S2(B)). The reduction in Moran’s *I* from raw diagnosed diabetes prevalence to OLS residuals (0.5734 to 0.1696) suggests that physical inactivity, low food access, and border-region status captured a substantial portion of the observed spatial patterning across Texas counties. However, the remaining significant residual spatial autocorrelation indicates that additional unmeasured spatially structured factors may still influence county-level diabetes burden. This residual spatial dependence is acknowledged as a limitation; future analyses incorporating spatial error or spatial lag models are recommended. Although residual spatial autocorrelation remained statistically significant, the substantial reduction in Moran’s *I* from the outcome variable to the OLS residuals indicates that the included predictors captured a meaningful portion of the spatial patterning in county-level diabetes burden. For the present study objective, which was to estimate the adjusted association between La Paz border-region status and diagnosed diabetes prevalence, the OLS framework with HC3 robust standard errors provided a transparent and interpretable baseline model. However, because HC3 standard errors do not directly model spatial dependence, these findings should be interpreted as ecological associations, and future analyses using spatial error, spatial lag, or geographically weighted regression models are warranted.

## 4 Discussion

This cross-sectional ecological analysis of 253 Texas counties shows that border-region county status was significantly associated with diagnosed diabetes prevalence after adjustment for physical inactivity and low food access. To our knowledge, this is among the first studies to evaluate this relationship using the official La Paz Agreement border definition, VIF-corrected regression with HC3 robust standard errors, formal spatial autocorrelation testing, and current federal data across Texas counties.

Border-region counties had 33% higher mean diagnosed diabetes prevalence in unadjusted descriptive comparisons (16.1% vs. 12.1%). After adjustment for physical inactivity and low food access, border-region status remained associated with a 0.625 percentage-point higher diagnosed diabetes prevalence. The choropleth map (Fig. 3) shows that this elevated burden is geographically concentrated along the Texas–Mexico border region, a pattern confirmed formally by Moran’s *I* (*I* = 0.5734, *p* = 0.001).

The persistence of the border-region association (*β* = 0.625, *p <* 0.001) after controlling for physical inactivity and low food access suggests that structural and geographic disadvantages unique to border communities may contribute to elevated diabetes burden beyond the included behavioral and food-access covariates. Such factors may include limited primary care capacity and specialist access in medically underserved areas [17, 23], occupational exposures among agricultural and manual labor workforces [2], high rates of undiagnosed diabetes and barriers to early detection [24, 25], historical underinvestment in health infrastructure [6], transportation barriers that restrict access to both healthcare and nutritious food, and cultural and linguistic barriers to healthcare engagement [8].

The finding that low food access alone was not a significant independent predictor, while compounded food and income disadvantage was significantly correlated with diagnosed diabetes prevalence, adds nuance to the food environment literature [13, 14, 12]. This suggests that food environment interventions in isolation may be insufficient. Instead, diabetes prevention strategies in border-region counties may require integrated approaches that address food availability, household income, transportation access, healthcare access, and culturally responsive prevention programs.

The observed associations, despite the temporal gap in food-environment data, underscore the potential long-term influence of structural food-access conditions on chronic disease burden. The 2015 food-access measure may reflect historical built environment conditions that preceded the diagnosed diabetes patterns observed in 2025, consistent with the slow-evolving nature of grocery-store infrastructure, transportation access, and neighborhood-level resource distribution.

The high model fit observed in the regression analysis should be interpreted in light of the ecological and model-based data structure. Diagnosed diabetes prevalence and physical inactivity were both obtained from the CDC PLACES multilevel regression and poststratification framework. Therefore, the high coefficient of determination (*R*^2^ = 0.960) reflects strong structural alignment among modeled county-level health indicators rather than individual-level predictive certainty. This interpretation is consistent with the study design, which aimed to quantify ecological associations across Texas counties rather than infer individual-level risk.

The reduction in Moran’s *I* from raw diagnosed diabetes prevalence to OLS residuals (0.5734 to 0.1696) suggests that the model predictors captured a substantial portion of the observed spatial patterning across Texas counties. However, the remaining significant residual spatial autocorrelation (*I* = 0.1696, *p* = 0.001) indicates that neighboring counties still share similar unexplained diabetes burden after adjustment for physical inactivity, low food access, and border-region status. This remaining spatial dependence may reflect unmeasured spatially structured factors, such as shared healthcare infrastructure, regional economic conditions, insurance coverage, transportation networks, or cultural dietary patterns. Although spatial error, spatial lag, or geographically weighted regression models would be useful extensions, the current OLS framework with HC3 robust standard errors provides a transparent and interpretable baseline estimate of the adjusted border-region association.

The sensitivity analysis confirmed that the border-region association was significant under both geographic specifications, providing robustness evidence that the observed association was not solely dependent on the specific border definition used.

## 5 Limitations and Future Work

This study has several limitations. First, because the analysis was ecological and county-level, the observed associations may not reflect individual-level relationships. Therefore, the findings should not be interpreted causally. In addition, the cross-sectional design prevents assessment of temporal ordering between border-region status, food access, physical inactivity, and diagnosed diabetes prevalence.

Second, the food-access variables reflect 2015 data and may not fully capture current food environment conditions in Texas counties. However, food-access infrastructure is a structural determinant that may change gradually over time; therefore, these metrics remain a plausible proxy for the historical environmental context shaping current chronic disease patterns. Nevertheless, the temporal gap between the 2015 food-access measure and the 2025 health estimates remains an important limitation.

Third, diagnosed diabetes, obesity, and physical inactivity were all derived from the CDC PLACES multilevel regression and poststratification framework. This shared model-based origin may contribute to the high model fit (*R*^2^ = 0.960), because the OLS model identifies structured ecological alignment among pre-modeled county-level indicators. While this limits the ability to observe individual-level stochastic variation, it provides an interpretable assessment of ecological relationships between geographic status and modeled public health estimates.

Fourth, although the inclusion of physical inactivity, low food access, and border-region status substantially reduced spatial clustering, significant residual spatial autocorrelation remained in the OLS residuals (Moran’s *I* = 0.1696, *z* = 4.64, *p* = 0.001). This suggests that additional spatially structured factors may still influence county-level diabetes burden. Future studies should apply spatial error, spatial lag, or geographically weighted regression models to more directly account for geographic dependence among Texas counties. Additional covariates, including Hispanic ethnicity percentage, insurance coverage, healthcare utilization, poverty, rurality, and primary care availability, should also be incorporated. While residual spatial dependence remains, the current OLS approach with HC3 robust standard errors serves as an interpretable first-step model for quantifying the adjusted border-region association. This baseline estimate provides a clear reference point for future studies using more complex spatial econometric specifications.

Finally, because this study was limited to Texas, the findings may not generalize to other US–Mexico border states.

## 6 Conclusion

Border-region county status was significantly associated with diagnosed diabetes prevalence across Texas counties. Border-region counties showed 33% higher mean diagnosed diabetes prevalence in unadjusted comparisons and a statistically significant 0.625 percentage-point higher prevalence after adjustment for physical inactivity and low food access. Physical inactivity emerged as the strongest independent predictor.

Formal spatial autocorrelation testing confirmed strong geographic clustering of diagnosed diabetes prevalence (*I* = 0.5734, *p* = 0.001), which was substantially reduced in OLS residuals (*I* = 0.1696, *p* = 0.001). This reduction suggests that the model predictors captured a substantial portion of the observed spatial patterning, although significant residual spatial autocorrelation remained.

These findings suggest that structural and geographic disadvantages unique to border communities may contribute to elevated diabetes burden beyond lifestyle factors. The results support targeted public health investment in the Texas–Mexico border region, including expanded primary care infrastructure, diabetes prevention programs, physical activity promotion, transportation access improvements, and culturally responsive community health interventions.

## Ethics Statement

This study used only publicly available, de-identified county-level aggregate data from federal agencies (CDC and USDA). No individual-level human subjects data were collected or used. Institutional review board approval was not required.

## Data Availability

All primary data used in this analysis are publicly available from CDC PLACES (https://www.cdc.gov/places; county-level data available through https://data.cdc.gov) and the USDA Economic Research Service Food Environment Atlas (https://www.ers.usda.gov/data-products/food-environment-atlas). The processed county-level analytical dataset used for the final regression, correlation, descriptive, sensitivity, and spatial autocorrelation analyses has been deposited in Zenodo, together with the Python analysis code and figure-generation scripts: https://doi.org/10.5281/zenodo.20390172. No restricted, proprietary, or individual-level human subjects data were used in this study.

## Author Contributions

Conceptualization: P.R.S., S.K., Y.Y., M.A.A.M.

Data curation: P.R.S., M.A.A.M.

Formal analysis: Y.Y., P.R.S.

Methodology: Y.Y., P.R.S., S.K.

Software: Y.Y.

Visualization: Y.Y.

Writing – original draft: P.R.S., Y.Y.

Writing – review & editing: P.R.S., S.K., Y.Y., M.A.A.M.

## Acknowledgments

The authors acknowledge the Centers for Disease Control and Prevention and the US Department of Agriculture Economic Research Service for making publicly available the data used in this analysis. The authors also acknowledge the UTRGV high-performance computing resources used in this analysis.

## Supporting Information

**S1 File. Supplementary materials for “Border-Region Status and Diagnosed Diabetes Prevalence in Texas: A Cross-Sectional Ecological Analysis.”** This file includes OLS regression diagnostic plots, Global Moran’s *I* scatter plots, variance inflation factor analysis, sensitivity analysis using alternative border definitions, spatial autocorrelation summary, the complete La Paz Agreement border-county list, dataset description, and code availability statement.

